# Cytokeratin-18 has potential as a biomarker of drug-induced liver injury in European and African patients on treatment for tuberculosis

**DOI:** 10.1101/2020.06.03.20121038

**Authors:** Sarah AE Rupprechter, Derek J Sloan, Wilna Oosthuyzen, Till T Bachmann, Adam T Hill, Kevin Dhaliwal, Kate Templeton, Joshua Matovu, Christine Sekaggya-Wiltshire, James W Dear

## Abstract

Patients on anti-tuberculosis (anti-TB) therapy are at risk of developing drug-induced liver injury (DILI). Cytokeratin-18 (K18) is an exploratory DILI biomarker that has been developed predominately in Caucasian populations and not African populations in whom TB is common. Our aim was to determine the K18 concentration in different populations with mycobacterial infection and investigate whether K18 has potential as a biomarker of anti-TB DILI.

European patients receiving anti-TB therapy were recruited at the Royal Infirmary of Edinburgh, UK (ALISTER - ClinicalTrials.gov Identifier: NCT03211208). African patients with HIV-TB coinfection, receiving anti-TB and anti-retroviral therapy (ART), were recruited at the Infectious Diseases Institute, Kampala, Uganda (SAEFRIF - NCT03982277). Serial blood samples, demographic and clinical data were collected. K18 was quantified using the M65 ELISA.

The study had 235 participants (healthy volunteers (n=28); ALISTER: active TB (n=30), latent TB (n=88), non-tuberculous mycobacterial infection (n=25); SAEFRIF: HIV-TB coinfection (n=64)). There was no difference in K18 across the groups and treatment did not affect K18 in the absence of DILI. Alanine transaminase activity (ALT) and K18 were correlated (r=0.42, 95%CI=0.34-0.49, P<0.0001). Variability was higher for K18 than ALT. There were two DILI cases: baseline ALT was 18 and 28 IU/1, peak ALT 431 and 194 IU/L; baseline K18 58 and 219 U/L, peak K18 1247 and 3490 U/L, respectively.

Circulating K18 was comparable in UK and Ugandan patients. K18 correlated with ALT and increased with DILI. Further work should determine the diagnostic and prognostic utility of K18 in this global context-of-use.

## Introduction

Tuberculosis (TB) is one of the top 10 global causes of death, with an estimated 10 million new cases, 1.2 million deaths among HIV-negative individuals and 251,000 deaths among HIV-positive individuals in 2018 (World Health Organization (WHO), 2019). The global burden of TB is unequally distributed, disproportionately affecting low- and middle-income countries particularly in Africa and Asia. One of the reasons for this is HIV-TB coinfection. Individuals with latent TB typically have a 5-10% chance of developing active TB over the course of their life, however, in HIV-infected individuals this risk rises to 10% annually (Corbett *et al*., 2003). HIV-TB coinfection is present in one third of all HIV-infected individuals and causes one quarter of HIV-related deaths (Getahun *et al*., 2010).

One of the barriers to the effective treatment of TB are the adverse drug reactions experienced by patients on anti-TB medications, with one of the most common being drug-induced liver injury (DILI) (Saukkonen *et al*., 2006). Three of the four first line drugs used in the treatment of TB - isoniazid, rifampicin and pyrazinamide - are potentially hepatotoxic (Girling, 1977). The estimate of the incidence of DILI in individuals undergoing anti-TB treatment for active TB varies from 2 to 33% depending on the cohort studied, drug regimen used, monitoring and reporting practices (Saukkonen *et al*., 2006; Tostmann *et al*., 2008; Ramappa and Aithal, 2013). Individuals who experience DILI often need to stop treatment and, if clinically indicated, recommence once liver function tests (LFTs) return to normal. However, for some individuals re-exposure to the same drugs leads to reoccurrence of DILI (Metushi, Uetrecht and Phillips, 2016), and for others liver injury progresses even after treatment has stopped (Hassan *et al*., 2015).

Diagnosis of DILI relies on LFTs, with alanine aminotransferase (ALT) activity considered one of the gold standards for determining liver injury. The DILI Expert Working Group defines DILI as ≥ 3 × upper limit of normal (ULN) of ALT in the presence of symptoms, or ≥ 5 × ULN ALT in the absence of symptoms (Aithal *et al*., 2011). Although ALT is currently the gold standard for determining DILI there are issues associated with its use. ALT is not specific to the liver and can provide false positive results, with elevations in ALT occurring after muscular damage following exercise (Pettersson *et al*., 2008) or subsequent to a myocardial infarction (LaDue and Wroblewski, 1955). Furthermore, elevations in ALT are not specific to DILI (Senior, 2012) and can occur due to metabolic perturbations (Sattar *et al*., 2004)(Hanley *et al*., 2004). In paracetamol-overdose DILI there is a delay between insult to the liver and rise in ALT, (Antoine and Dear, 2016), meaning ALT is not optimal as a biomarker of DILI in this context. To address these challenges recent work has identified novel biomarkers capable of diagnosing, and in some cases predicting, DILI with more accuracy.

Cytokeratin-18 (K18) is an intermediate filament protein responsible for maintaining the cytoskeletal structure in the liver and other epithelial cells and is reported to make up 5% of the liver’s total protein content (Adebayo, Mookerjee and Jalan, 2012). K18 is a mechanistic biomarker of liver injury, providing information on the pattern of cell death. In apoptosis, the release of a caspase-cleaved form of K18 (CC-K18) is an early event during cellular structural rearrangement (Caulín, Salvesen and Oshima, 1997). Cleavage of K18 by caspases 3, 6 and 7 generates fragments of 30 kDa and 45 kDa (Eguchi, Wree and Feldstein, 2014). Whereas, in necrosis, the full-length form of K18 (FL-K18) is passively released upon cell death (Caulín, Salvesen and Oshima, 1997). Both FL-K18 and CC-K18 have been used to quantify the pattern of hepatocyte death in the liver diseases including non-alcoholic steatohepatitis (NASH) (Wieckowska *et al*., 2006) and hepatitis C (Bantel *et al*., 2004). A recent multi-centre study has shown that K18 can predict acute liver injury after paracetamol overdose at hospital presentation and has superior sensitivity compared with ALT (Dear *et al*., 2018). K18 has the potential to aid the process of preclinical and clinical drug development and this is acknowledged by regulatory support from the US Food and Drug Administration and the European Medicines Agency (European Medicines Agency, 2016; Food and Drug Administration Centre for Drug Evaluation and Research, 2016). A recent study from the Critical Path Institute’s Predictive Safety Testing Consortium (PSTC) in the United States and the European Safer and Faster Evidence based Translation (SAFE-T) consortium has determined the normal healthy reference interval for FL-K18 as 0-151.14 U/L and for CC-K18 52.46-373.55 U/L (Church *et al*., 2018). Although this important paper has defined a healthy reference interval, there are still limited data on the circulating concentration of K18 in other populations, including in Africans.

The aim of this study was to determine the K18 concentration in different populations with mycobacterial infections and investigate whether K18 has potential as a biomarker of anti-TB DILI. As the liver is commonly affected by mycobacterial infection it is essential to determine the effect of mycobacterial infection on K18 in the absence of DILI. Therefore, the K18 concentration was determined in UK and African patients receiving anti-TB treatment and healthy volunteers. The effect of starting treatment, the variability of K18 and the correlation of K18 with ALT was determined.

## Methods

The participants in this study came from three different studies. Healthy volunteers were recruited into the “MicroRNA signatures of disease activity in ANCA-associated vasculitis” study at the Royal Infirmary, Edinburgh. Participants with active TB, latent TB and non-tuberculous mycobacterial infection were recruited into the “Assessing Antibiotic-Induced Liver Injury for the Stratification of Tuberculosis Patients” (ALISTER) clinical study at the Royal Infirmary, Edinburgh (ClinicalTrials.gov Identifier: NCT03211208). Participants with HIV-TB coinfection were recruited into the “Safety and Efficacy of High Dose Rifampicin in Tuberculosis (TB)-HIV Co-infected Patients on Efavirenz- or Dolutegravir-based Antiretroviral Therapy” (SAEFRIF) clinical trial at the Infectious Disease Institute, Kampala, Uganda (ClinicalTrials.gov Identifier: NCT03982277).

## ALISTER clinical study

Participants were recruited at the Royal Infirmary of Edinburgh. Adults (≥16 years, ≤85 years), receiving treatment for active or latent tuberculosis, or non-tuberculous mycobacterial infection were included. Patients were excluded if they did not have the capacity to provide informed consent or were known to be HIV positive. Full written informed consent was obtained from every participant and the study was approved by the West of Scotland Research Ethics Committee.

Patients were classified as having active tuberculosis either if they had culture confirmation of *M. tuberculosis* and presence of active disease, or if a clinician decided there was sufficient evidence of active disease to start them on treatment. Latent tuberculosis patients had a positive interferon-gamma release assay (IGRA) and no evidence of active disease. Patients with non-tuberculous mycobacterial (NTM) infection had grown at least 2 cultures with non-tuberculous mycobacterium, had clinical signs of pulmonary disease and an exclusion of other relevant diagnoses (Griffith *et al*., 2007; Haworth *et al*., 2017).

Active and latent TB patients were treated following WHO guidelines (World Health Organization (WHO), 2017, 2018). Patients with susceptible active TB were treated with isoniazid, rifampicin, ethambutol and pyrazinamide for an initiation phase of 2 months, followed by rifampicin and isoniazid for a continuation phase of 4 months (World Health Organization (WHO), 2017). Patients with latent TB were treated with either a combination of isoniazid and rifampicin for 3 months, or isoniazid or rifampicin alone for 6 months (World Health Organization (WHO), 2018). Patients with *M. avium* complex (MAC) infection were treated with the recommended regimen of rifampicin, ethambutol and clarithromycin for 2 years (Haworth *et al*., 2017). Dependent on the NTM species, disease severity, resistance profile of the infection and tolerance of the individual, drugs were replaced or added, including isoniazid, moxifloxacin, azithromycin and amikacin (Griffith *et al*., 2007; Haworth *et al*., 2017).

## SAEFRIF clinical trial

The “Safety and Efficacy of High Dose Rifampicin in Tuberculosis (TB)-HIV Co-infected Patients on Efavirenz- or Dolutegravir-based Antiretroviral Therapy” (SAEFRIF) clinical trial (NCT03982277) is currently running at the Infectious Disease Institute at Makerere University, Uganda. The data presented in this paper provide preliminary results of the trial. This trial was approved by a local research and ethics committee, the National Drug Authority and the Uganda National Council for Science and Technology. The study was performed in accordance with the general principles in the International Ethical Guidelines for Biomedical Research Involving Human Subjects and the Declaration of Helsinki. Inclusion criteria were patients aged ≥18 years who had confirmed HIV-1 infection, were already on efavirenz (EFV)-based or dolutegravir (DTG)-based ART or planning to start ART and were diagnosed with TB and due to initiate rifampicin-containing therapy. Further inclusion criteria included providing informed consent and complying with scheduled trial visits, treatment, laboratory tests and other study procedures. Exclusion criteria were patients who have rifampicin resistant TB, pregnant women or those planning on getting pregnant during treatment, women of reproductive age on DTG who decline the use of effective contraceptive, patients with liver disease, ALT > x5 ULN or glomerular filtration rate < 50 ml/min.

The protocol for this trial is described in Nabisere *et al*., 2020. Patients were randomised to one of 4 arms of the trial for the initiation period of treatment, either the standard dose of rifampicin (10 mg/kg) or high dose rifampicin (35 mg/kg), along with either efavirenz- or dolutegravir-based ART. In addition, patients received standard TB therapy consisting of isoniazid, pyrazinamide and ethambutol for the initiation period of 2 months. Baseline blood samples (serum) were collected before commencing treatment and further samples were collected at weeks 2, 4, 6, and 8 of treatment.

### Data and blood samples

For the ALISTER study and SAEFRIF trial demographic and clinical data from the participants were recorded from medical records and clinical trial records. Liver function test results (ALT) were recorded from each clinic visit Blood samples (serum (SAEFRIF) and plasma (ALISTER)) were collected at first clinic visit and subsequent clinic visits. Once collected blood was processed by centrifugation, then the supernatant was aliquoted and stored at -80°C.

### Quantification of K18

Samples were stored at −80°C before analysis. The period of storage was (median 27.1; IQR 16.6-46.3; range 2.6-110.6 weeks). Total K18 was quantified using the Peviva M65 classic ELISA (Bioaxxes, Tewkesbury, UK), according to the manufacturer’s instructions. Samples were measured in duplicate. CV values for the dataset were (median [IQR]: 2.85%, [1.33-4.97%]).

### Assessment of causality

DILI was pre-defined in this study as >x3 ULN ALT in the presence of symptoms or >x5 ULN in absence of symptoms (Aithal *et al*., 2011). The ULN for ALT was 50IU/L in the populations studied. The Roussel Uclaf Causality Assessment Method (RUCAM) was used to determine formal causality between anti-TB medication and liver injury. The pattern of liver injury was determined using the R ratio, considering ALT and alkaline phosphatase activity. Further factors considered include time to onset, course of injury, risk factors (age and alcohol), concomitant drugs, the exclusion on non-drug causes of injury and previous information on drug hepatotoxicity (Bethseda (MD): National Institute of Diabetes and Digestive and Kidney Diseases, 2017).

### Statistical analysis

Data were summarised as median (IQR) or n (%) for summary statistics of the study participants. One-way Kruskal-Wallis ANOVA was used to determine the difference in K18 between the different patient groups. Wilcoxon matched-pairs signed rank test was used to determine the difference in K18 upon starting treatment. The coefficient of variation was calculated across 3 or more samples to assess the intra-individual variability of ALT and K18 in the HIV-TB coinfected population. Correlation of ALT and K18 was determined using Spearman’s rank correlation. The difference between K18 in samples grouped by ALT was determined using a Mann-Whitney t-test. Statistical analyses were performed using Graphpad Prism (GraphPad Software, La Jolla, California).

## Results

A total of 235 patients were recruited into the study, (healthy volunteers (n=28). In ALISTER: active TB (n=30), latent TB (n=88), non-tuberculous mycobacterial infection (n=25). In SAEFRIF: HIV-TB coinfection (n=64)). Demographic and clinical characteristics are presented in **Table 1**.

**Table 1:** Clinical and demographic characteristics of patients in the study. Data are presented as median (IQR) or n (%).

|  | Median (IQR) N (%) | Active TB<br>(n=30) | Latent<br>TB<br>(n=88) | NTM<br>infection<br>(n=25) | HIV-TB<br>coinfection<br>(n=64) |
| --- | --- | --- | --- | --- | --- |
| **Age (yrs)** |  | 41<br>(33-56) | 40<br>(23-63) | 66<br>(57-74) | 38<br>(32-44) |
| **Gender** | **Male** | 20 (67%) | 23 (26%) | 10 (40%) | 40 (62%) |
|  | **Female** | 10 (33%) | 65 (74%) | 15 (60%) | 24 (38%) |
| **Ethnicity** | **African** | 2 (7%) | 7 (8%) | 0 (0%) | 64 (100%) |
|  | **Arab** | 0 (0%) | 1 (1%) | 0 (0%) | 0 (0%) |
|  | **Bangladeshi** | 1 (3%) | 2 (2%) | 0 (0%) | 0 (0%) |
|  | **Indian** | 7 (23%) | 3 (3%) | 0 (0%) | 0 (0%) |
|  | **Iraqi** | 1 (3%) | 0 (0%) | 0 (0%) | 0 (0%) |
|  | **Pakistani** | 5 (17%) | 3 (3%) | 1 (4%) | 0 (0%) |
|  | **South East Asian** | 2 (7%) | 1 (1%) | 0 (0%) | 0 (0%) |
|  | **White** | 12 (40%) | 66 (75%) | 24 (96%) | 0 (0%) |
|  | **Unknown** | 0 (0%) | 5 (6%) | 0 (0%) | 0 (0%) |
| **Location of infection** | **Pulmonary** | 11 (37%) | - | - | 51 (80%) |
|  | **Extrapulmonary** | 16 (53%) | - | - | 9 (14%) |
|  | **Both** | 3 (10%) | - | - | 1 (1%) |
|  | **Unknown** | 0 (0%) | - | - | 3 (5%) |
| **Culture confirmed** | **Yes** | 20 (67%) | - | - | 64 (100%) |
|  | **No** | 10 (33%) | - | - | 0 (0%) |
| **Resistance** | **None** | 26 (87%) | - | - | - |
|  | **Isoniazid** | 1 (3%) | - | - | - |
|  | **Pyrazinamide** | 2 (7%) | - | - | - |
|  | **Rifampicin** | 0 (0%) | - | - | - |
|  | **MDR** | 1 (3%) | - | - | - |
| **NTM species** | **MAC** | - | - | 22 (88%) | - |
|  | **M. abscessus** | - | - | 2 (8%) | - |
|  | **M. malmoense** | - | - | 1 (4%) | - |
| **Baseline ALT (IU/L)** |  | 18<br>(14-35) | 15<br>(12-21) | 15<br>(12-22) | 20<br>(14-30) |
| **Treatment** | **Isoniazid, Rifampicin** | - | 43 (48%) | - | - |
|  | **Isoniazid** | - | 26 (30%) | - | - |
|  | **Rifampicin** | - | 18 (20%) | - | - |
|  | **Moxifloxacin** | - | 1 (1%) | - | - |
|  | **Rifampicin,**<br>**Azithromycin** |  | - | 1 (4%) | - |
|  | **Rifampicin,**<br>**Clarithromycin** | - | - | 3 (12%) | - |
|  | **Rifampicin,**<br>**Clarithromycin,**<br>**Amikacin** | - | - | 1 (4%) | - |
|  | **Rifampicin,**<br>**Ethambutol** | - | - | 1 (4%) | - |
|  | **Rifampicin,**<br>**Ethambutol, Amikacin** | - | - | 1 (4%) | - |
|  | **Rifampicin,**<br>**Ethambutol,**<br>**Clarithromycin** | - | - | 15 (60%) | - |
|  | **Rifampicin,**<br>**Ethambutol,**<br>**Moxifloxacin** | - | - | 1 (4%) | - |
|  | **Rifabutin,**<br>**Clarithromycin,**<br>**Moxifloxacin** | - | - | 1 (4%) | - |
|  | **Clarithromycin,**<br>**Clofazimine,**<br>**Azithromycin** | - | - | 1 (4%) | - |
|  | **Isoniazid, Rifampicin,**<br>**Pyrazinamide,**<br>**Ethambutol plus ART** | - | - | - | 64 (100%) |
| **Initiation phase** | **Isoniazid, Rifampicin,**<br>**Pyrazinamide,**<br>**Ethambutol** | 22 (73%) | - | - | - |
|  | **Isoniazid, Rifampicin,**<br>**Pyrazinamide,**<br>**Moxifloxacin** | 1 (3%) | - | - | - |
|  | **Isoniazid, Rifampicin,**<br>**Pyrazinamide,**<br>**Ethambutol,**<br>**Moxifloxacin** | 1 (3%) | - | - | - |
|  | **Isoniazid, Rifabutin,**<br>**Pyrazinamide,** | 1 (3%) | - | - | - |
|  | **Ethambutol** |  |  |  |  |
|  | **Isoniazid, Rifampicin, Ethambutol, Moxifloxacin** | 3 (10%) | - | - | - |
|  | **Rifampicin, Ethambutol, Moxifloxacin** | 1 (3%) | - | - | - |
|  | **Bedaquiline, Clofazimine, Cycloserine** | 1 (3%) | - | - | - |
| **Continuation phase** | **Isoniazid, Rifampicin** | 20 (67%) | - | - | - |
|  | **Isoniazid, Rifampicin, Moxifloxacin** | 2 (7%) | - | - | - |
|  | **Isoniazid, Rifampicin, Ethambutol** | 2 (7%) | - | - | - |
|  | **Isoniazid, Rifabutin, Moxifloxacin** | 1 (3%) | - | - | - |
|  | **Isoniazid, Ethambutol, Moxifloxacin** | 1 (3%) | - | - | - |
|  | **Isoniazid, Rifampicin, Pyrazinamide, Ethambutol** | 1 (3%) | - | - | - |
|  | **Rifampicin, Ethambutol, Moxifloxacin** | 2 (7%) | - | - | - |
|  | **Bedaquiline, Clofazimine, Cycloserine** | 1 (3%) | - | - | - |

Circulating K18 concentration and ALT activity was measured in healthy volunteers and samples collected from patients in the ALISTER study and SAEFRIF trial, **Figure 1**. There was no difference in K18 between groups (median [IQR]: healthy volunteers 211 [134-259]; active TB 154 [101-267]; latent TB 152 [106-218]; NTM infection 163 [138-263]; HIV-TB coinfection 178 [133-324] U/L). Serial samples were collected in 115 patients (ALISTER n=65 **(Figure 2);** SAEFRIF n=50 **(Figure 3))**. Starting treatment did not lead to a significant increase in ALT activity (median [IQR]: baseline 18 [13-27]; on treatment 20 [14-29] IU/L; P=0.06). Furthermore, there was no significant change in K18 concentration (median [IQR]: baseline 171 [113-267]; on treatment 167 [115-251] U/L; P=0.8).

**Figure 1:**
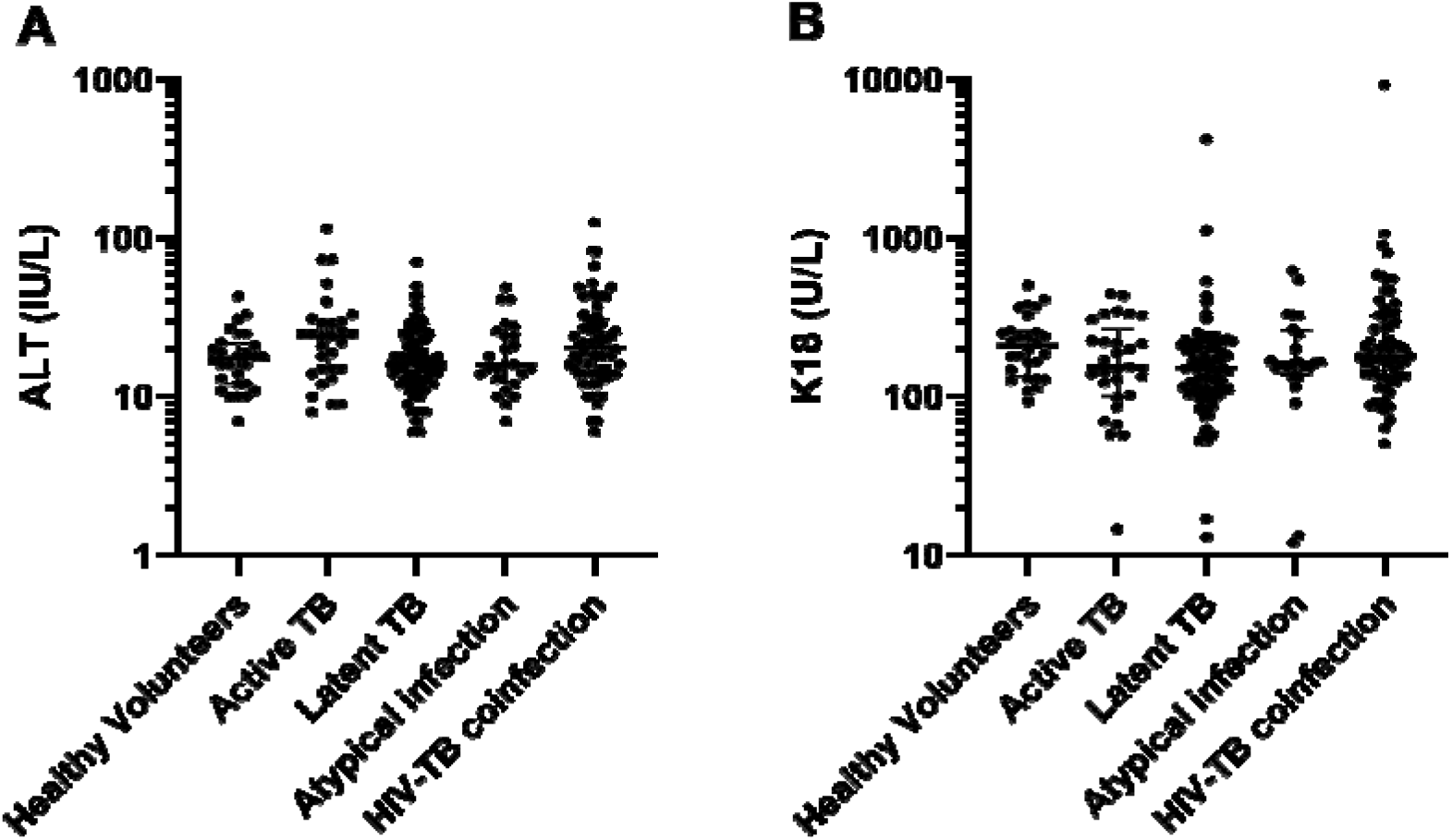
Circulating concentration of ALT (A) and K18 (B) in the first collected samples from the ALISTER study or SAEFRIF trial. Data are presented as dot plots. Line shows median and bars show interquartile range. Participants include healthy volunteers (n=28), active TB (n=30), latent TB (n=88), NTM infection (n=25) and HIV-TB coinfection (n=64).

**Figure 2:**
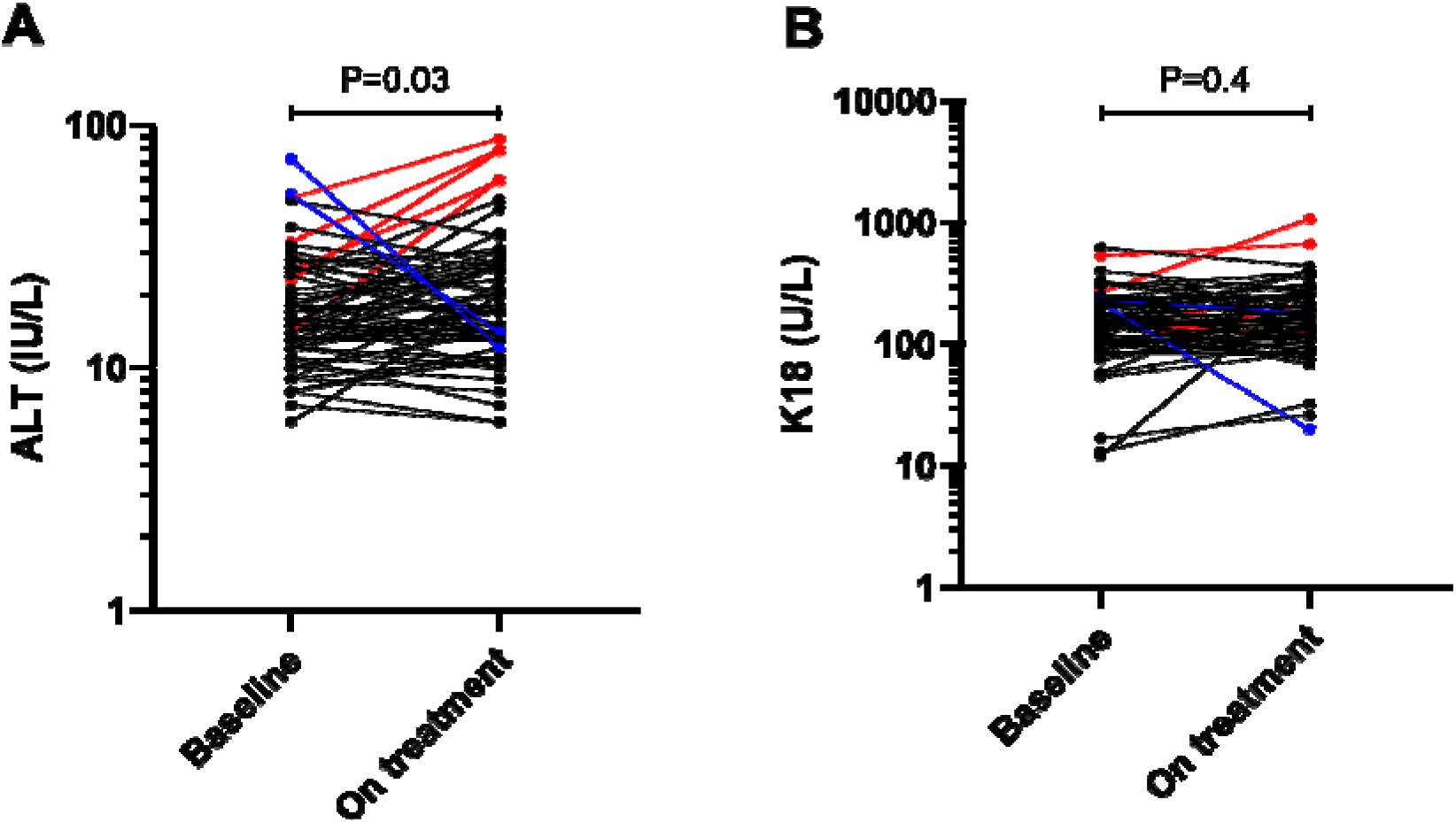
Circulating concentration of (A) ALT (IU/L) and (B) K18 (U/L) in sequential samples in patients within the ALISTER study, (active TB n=9; latent TB n=46; NTM n=10) Data shown as dot plots. Black dots show patients with normal ALT activity at baseline and on treatment; blue dots show patients with ALT activity >50U/L at baseline which decreased on treatment; red dots show patients whose ALT increased above 50U/L with treatment. The significance of differences between baseline and on treatment concentrations of biomarkers was determined by Wilcoxon signed rank test.

**Figure 3:**
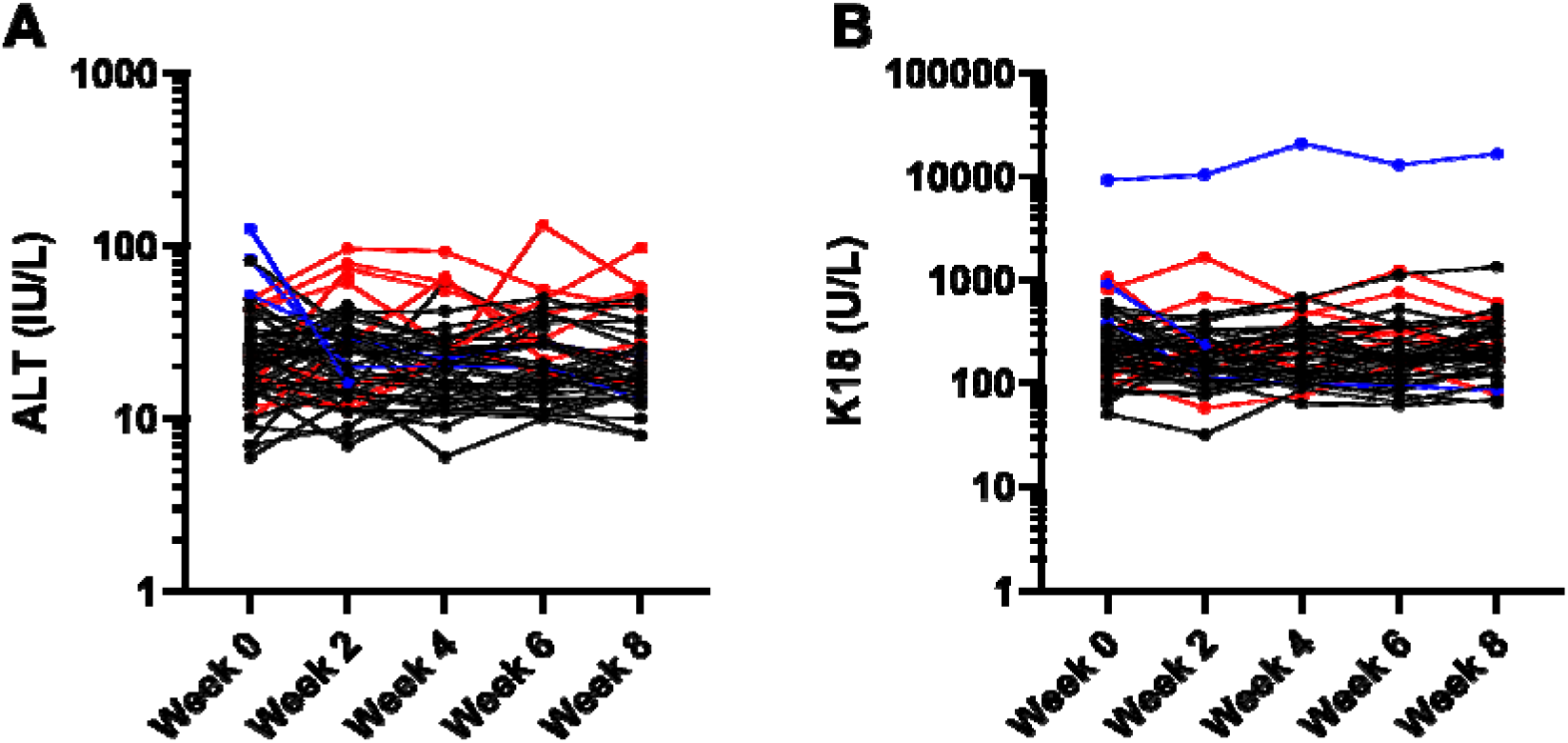
Circulating concentration of (A) ALT (IU/L) and (B) K18 for sequential samples in patients within the SAEFRIF trial. Black dots show patients with normal ALT throughout treatment, red points show patients whose ALT rises >50 IU/L during treatment, blue points show patients whose ALT falls from >50 IU/L upon starting treatment.

When all samples from all time points were included there was a significant correlation between ALT and K18 (N=491, Spearman rank r = 0.42, 95% CI = 0.34-0.49, P<0.0001) **(Figure 4A)**. K18 was increased 2.3 fold in those samples with elevated ALT (>50 IU/L) compared to samples with ALT<50 IU/L (median [IQR]: >50, 395 [217-683]; <50, 170 [120-250] U/L) **(Figure 4B)**. The inter-individual variability in K18 and ALT was compared in the different patient groups **(Table 2)**. Sequential samples from the SAEFRIF trial (patients with normal ALT (<50IU/L) and three or more samples collected) were analysed to determine the intra-individual variability over time (CV median [IQR]: ALT 23.9 [16.6-36.1] %; K18 35.4 [24.9-41.9] %; P=0.02). Inter- and intra-individual variability was higher for K18 than ALT.

**Figure 4:**
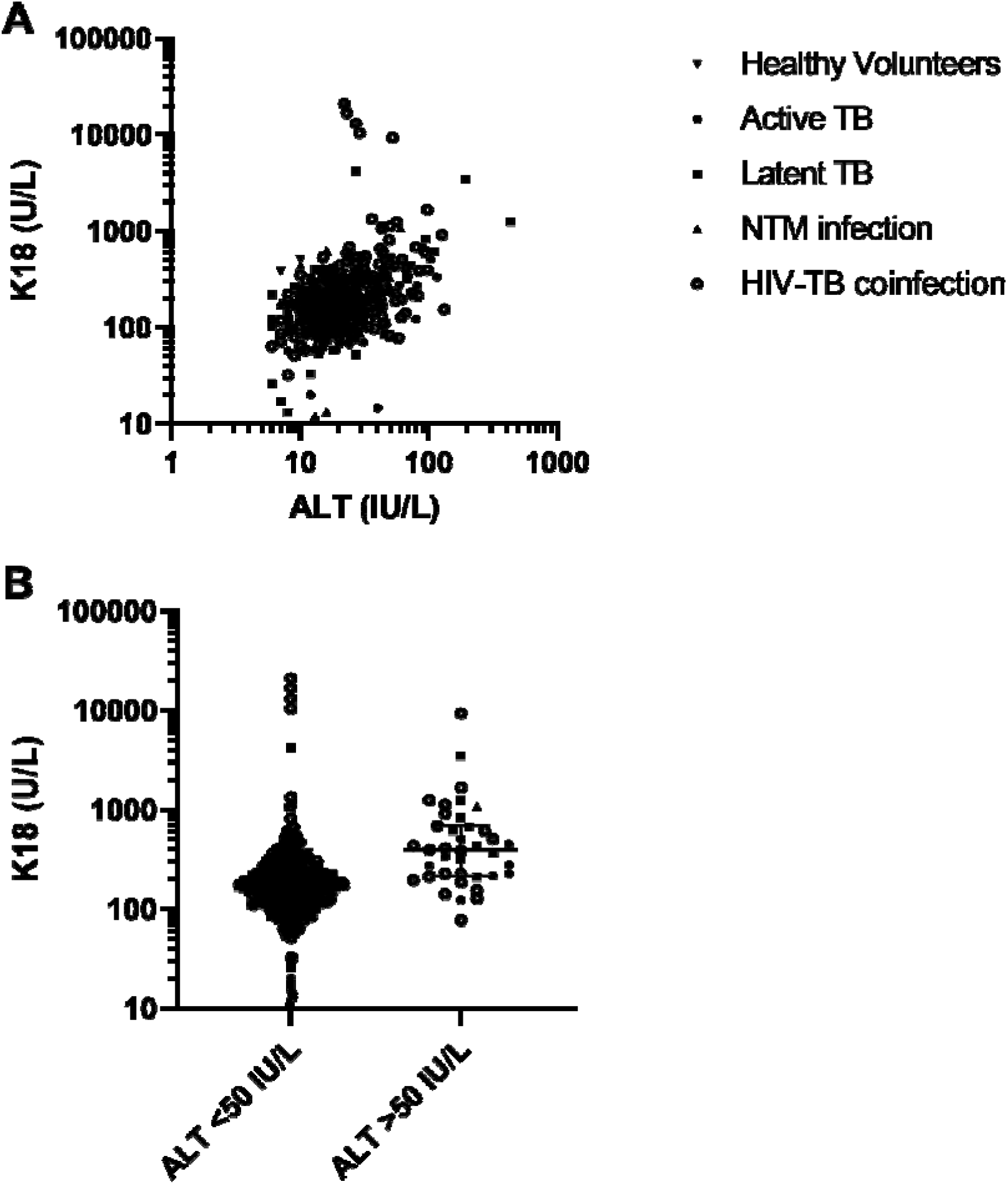
(A) Correlation of K18 (U/L) with ALT (IU/L). Statistical analysis of the significance of the correlation calculated using Spearman’s rank correlation coefficient (N=491, Spearman rank r = 0.42, 95% CI = 0.34-0.49, P<0.0001). (B) All samples grouped by normal ALT (<50 IU/L) (n=452) and elevated ALT (>50 IU/L) (n=39). The significance of the difference between the two groups was determined by Mann-Whitney t-test (P<0.0001).

**Table 2:**
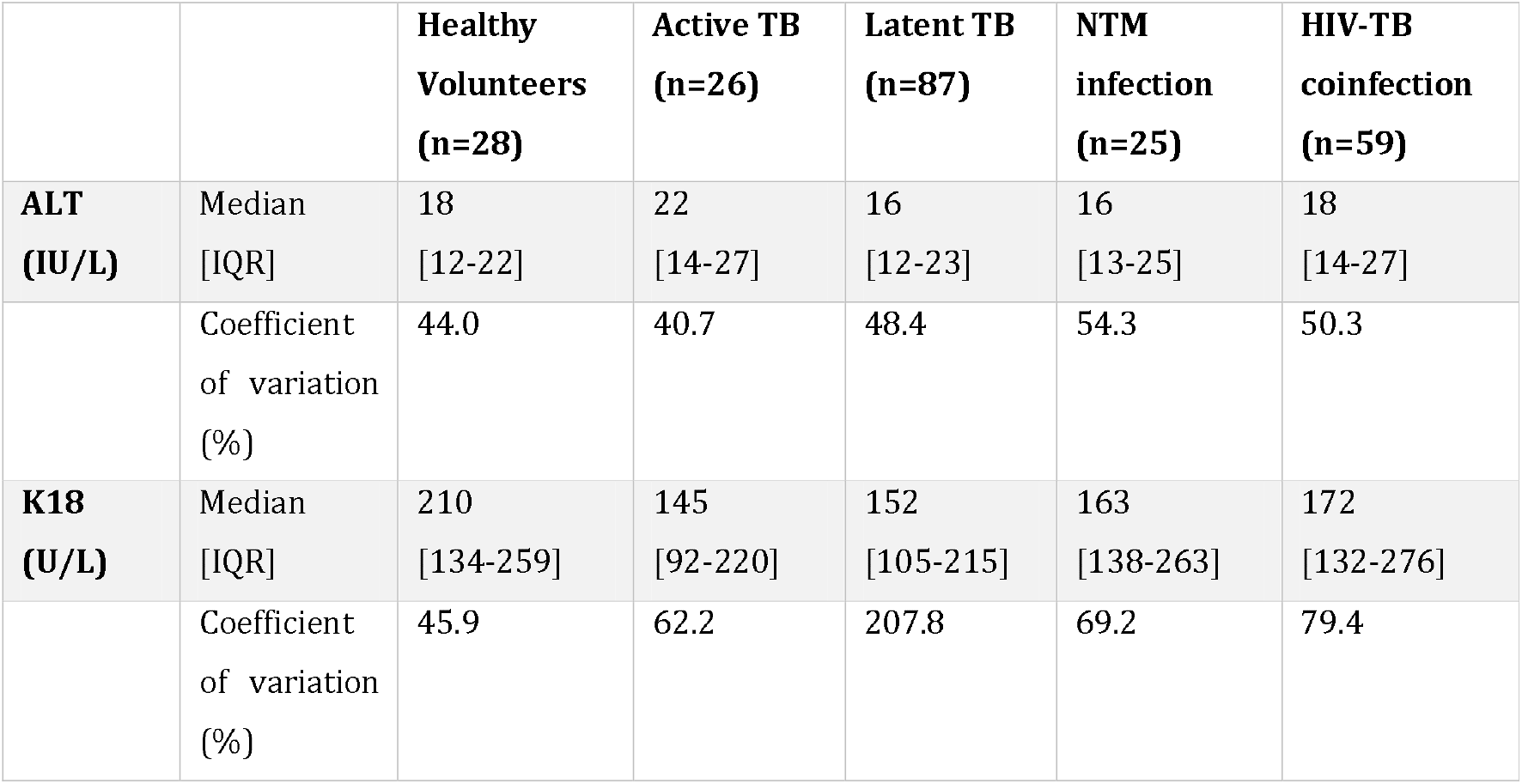
Circulating K18 and ALT in individuals with normal ALT (< 50 IU/L) in first samples taken upon starting ALISTER study or SAEFRIF trial.

|  |  | **Healthy**<br>**Volunteers**<br>**(n=28)** | **Active TB**<br>**(n=26)** | **Latent TB**<br>**(n=87)** | **NTM**<br>**infection**<br>**(n=25)** | **HIV-TB**<br>**coinfection**<br>**(n=59)** |
| --- | --- | --- | --- | --- | --- | --- |
| **ALT**<br>**(IU/L)** | Median<br>[IQR] | 18<br>[12-22] | 22<br>[14-27] | 16<br>[12-23] | 16<br>[13-25] | 18<br>[14-27] |
|  | Coefficient<br>of variation<br>(%) | 44.0 | 40.7 | 48.4 | 54.3 | 50.3 |
| **K18**<br>**(U/L)** | Median<br>[IQR] | 210<br>[134-259] | 145<br>[92-220] | 152<br>[105-215] | 163<br>[138-263] | 172<br>[132-276] |
|  | Coefficient<br>of variation<br>(%) | 45.9 | 62.2 | 207.8 | 69.2 | 79.4 |

In this study there were two cases of DILI (as pre-defined in study protocols), both cases were for patients receiving isoniazid alone for the treatment of latent TB within the ALISTER study. The first case was of a 51-year-old white Scottish male patient, who experienced peak ALT activity of 431 IU/L, **Figures 5A & B**. Before starting treatment, he had normal ALT (18 IU/L) and his K18 was 58 U/L. 3 months into treatment his ALT activity increased to 431 IU/L and K18 increased to 1248 U/L. Drug treatment was halted, ALT returned to within normal limits 18 weeks later. Formal causality between isoniazid and liver injury was determined using the RUCAM scale and was determined as probable (RUCAM scale = 6).

**Figure 5:**
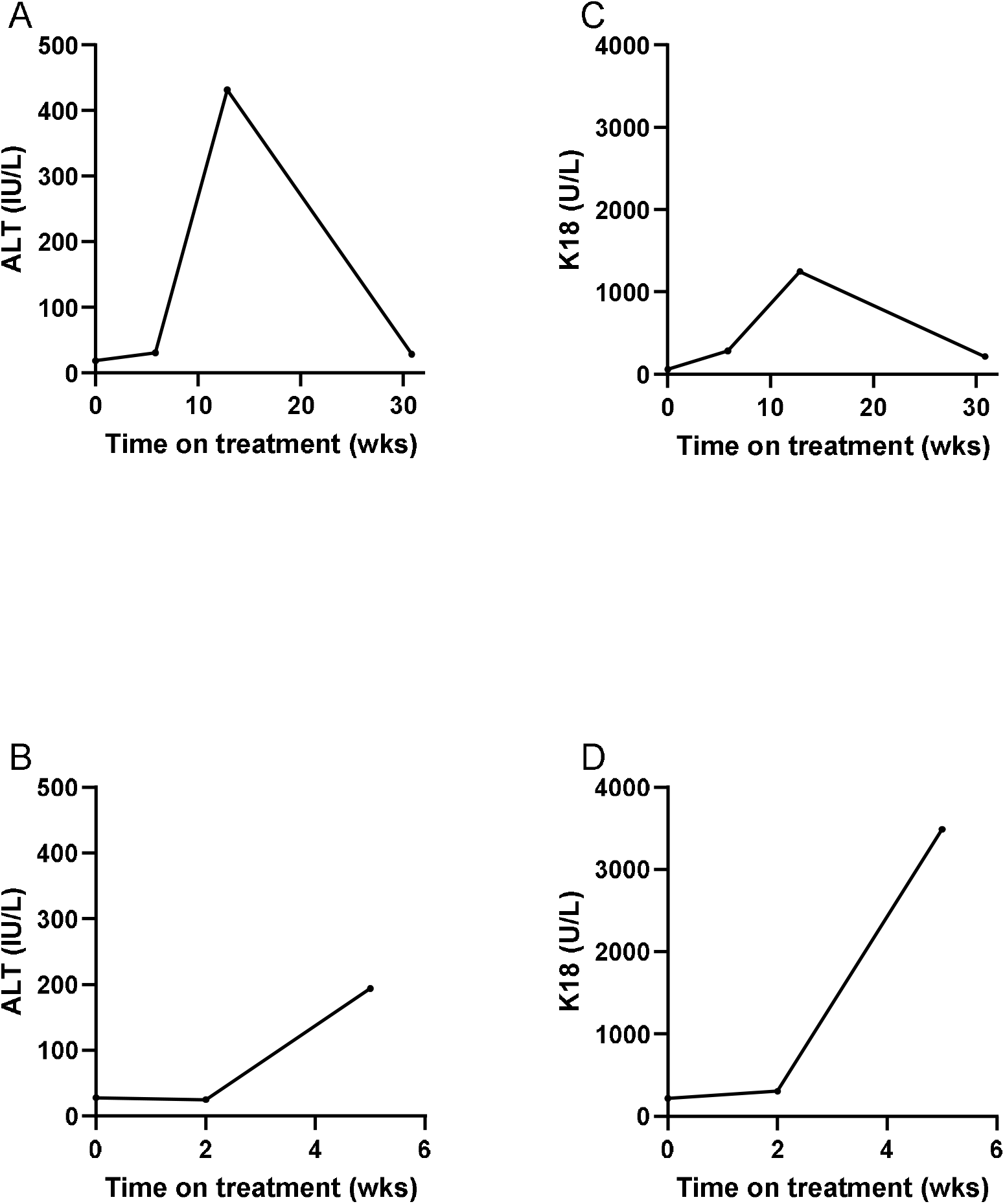
Circulating concentration of ALT [IU/L] (A & B] and K18 (U/L] (C & D] over the course of treatment [weeks] for two cases who developed DILI as pre-defined as >x3 ULN ALT in the presence of symptoms or >x5 ULN in absence of symptoms (Aithal *et al*., 2011]. Case 1 (A & C]; Case 2 (B & D].

The second case was of a 71-year-old white British female patient, **Figures 5C & D**. At baseline her ALT activity was 28 IU/L and K18 was 219 U/L. 2 weeks into treatment her ALT activity was still within normal range at 25 IU/L and K18 had risen to 307 U/L. 5 weeks into treatment DILI was determined with the presence of a drug rash and elevated ALT at 194 IU/L. At this time K18 activity had risen to 3490 U/L. RUCAM causality assessment indicated isoniazid was the probable cause of liver injury (RUCAM scale = 7).

## Discussion

In this study we have demonstrated that circulating K18 in healthy volunteers and patients with active TB, latent TB, non-tuberculous mycobacterial infection and HIV-TB coinfection are not significantly different. Furthermore, we have demonstrated that, in the absence of DILI, K18 does not change substantially upon commencing treatment K18 correlates with ALT. The intra and inter-patient variation of K18 was higher than that of ALT. Finally, in the two cases of DILI in this study, K18 increased substantially.

Several multicentre studies have demonstrated the utility of K18 as a biomarker of DILI in the context of predicting subsequent ALT activity increases after paracetamol overdose (Dear *et al*., 2018). These and other data from non-paracetamol DILI have resulted in K18 receiving FDA and EMA support as an exploratory DILI biomarker (European Medicines Agency, 2016). K18 has the potential to provides an insight into the mechanism of liver injury, with FL-K18 reporting cell death by necrosis and CC-K18 reporting cell death due to apoptosis.

Given the potential involvement of the liver in TB infection it was important to determine if circulating concentrations of K18 differed from healthy individuals in the presence of infection, including active, latent and non-tuberculous mycobacterial infection and HIV-TB coinfection. For example, if K18 was elevated by mycobacterial infection or HIV *per se* then it would be de-prioritised as a biomarker in this important context of use. In addition, there are limited data on K18 in different population groups, particularly Africans.

Our study indicates that the presence of active TB, latent TB, non-tuberculous mycobacterial infection and HIV-TB coinfection does not substantially affect circulating K18. Furthermore, patients in the SAEFRIF trial, who were African, had similar circulating K18 to those in the other groups, which were predominantly Caucasian. This suggests that the reference interval for an African and Caucasian population is likely to be similar. Patients with elevated ALT also had elevated K18, and ALT and K18 correlated significantly, indicating that K18 may have diagnostic utility. Within this study there were two cases of DILI, where elevations in ALT temporarily correlated with a rise in K18. The results of this study provide initial evidence for the potential use of K18 as a biomarker of TB medicine associated DILI. The variability of K18 was higher than ALT but not to the extent that this would be expected to limit clinical utility.

Further work should focus on determining the diagnostic value K18, whether it correlates with rises in ALT and so can diagnose DILI within this population. A clear definition of the dynamic range, sensitivity and specificity of K18 within this population is needed before it can be used as a biomarker of DILI. Furthermore, given that evidence suggests K18 rises earlier than ALT in paracetamol DILI, it is important to determine if K18 has the same predictive value in patients with mycobacterial infections. This predictive ability of K18 may enable early identification of patients at risk of DILI, leading to prevention of liver injury through halting or altering treatment regimens before significant liver injury develops. Specifically, K18 could be a useful early biomarker of the development of DILI in patients being reintroduced to essential anti-TB medications, a group at elevated risk of DILI recurrence. A limitation in translation to patient benefit is the lack of a point of care test for K18. Quantification of K18 currently relies on an ELISA which requires laboratory facilities and trained technicians. A cheap point of care test for K18 would enable testing in resource limited settings where TB infection is common.

One of the limiting factors for this study is the lack of an accurate reference interval for K18. A recent paper by Church et al., described the circulating concentrations of K18 in healthy controls and patients with DILI (Church *et al*., 2018). However, the ELISA used to perform the K18 analysis was different to the one used in this study. This makes comparison of the K18 data collected in this study to the published reference interval difficult In addition, the Church et al. study is limited by the populations used to generate the data, containing predominantly Caucasian individuals. Therefore, there is limited evidence of how these biomarkers perform in other populations. As a result, it is difficult to compare the data collected from HIV-TB coinfected Ugandans with the published healthy reference interval. Historical data suggested approximately 2-5% of patients receiving anti-TB treatment in the UK to develop DILI. However, within the ALISTER study only 2 patients developed DILI, 1.4% of the patients recruited. Larger multi-centre studies are required to recruit enough patients to determine the diagnostic power of K18 in anti-TB DILI.

In summary, the presence of TB infection does not alter K18 concentrations in the absence of DILI. Ugandan HIV-TB coinfected patients had similar K18 concentrations to healthy volunteers and Caucasian TB patients. Patients who experienced elevations in ALT also demonstrated rises in K18 indicating the diagnostic potential of K18. Future trials of K18 as a biomarker of anti-TB DILI could be performed using the data presented in this paper to inform the study design.

## Data Availability

Data will be available on request

